# Panel-level multilocus methylation quantification in native cell-free DNA by PCR-compatible sequential enzymatic processing

**DOI:** 10.64898/2026.06.11.26354897

**Authors:** Cintia Celina Vaquer, Paula Alida Wetten, Clara García Samartino, Jorge Daniel Rodriguez, Ricardo Nicolás Manzino, Roberto Pérez Ravier, Agustina Renata Angeloni, Rodrigo Damián Militello, Alejandro Ledesma, Juliana Sesma, Yanina Agnella, Erika Gudiño, Carina Evelyn Fracchia Díaz, Rodrigo Ongay, Guillermo Sanguinetti, Agustín Correa, Melani Carlen, Walter Ramón Minatti, Gastón Andrés Vaschalde, Paula Valdemoros, Leandro Sarrió, Luis Mayorga, Victoria Bocanegra, Emanuel Martín Campoy

## Abstract

DNA methylation provides informative signals for liquid biopsy, but low template abundance, distributed signals across loci and conversion-dependent processing can complicate implementation in low-input samples. Here we present Delta-HLD, a PCR-compatible platform that quantifies methylation directly in native DNA through sequential hybridization, ligation and methylation-sensitive digestion. By encoding locus specificity at the hemi-probe hybridization and ligation stage, Delta-HLD allows methylation-dependent products from multiple loci to converge on a shared amplification and detection system and contribute to a single panel-level readout. We established the assay chemistry, optimized panel composition through model-guided experiments and implemented the platform in qPCR and ddPCR, including a triplex qPCR format with per-sample digestion and amplification controls. Proof-of-concept plasma analyses showed discriminatory signals in colorectal cancer and supported transfer of the same assay logic to hepatocellular carcinoma. The assay was further adapted for cfDNA analysis following platelet-retaining processing, preserving cfDNA-associated signal while reducing genomic DNA recognition. Delta-HLD provides a compact native-DNA platform for multilocus methylation analysis in low-input settings without base conversion.

## Introduction

DNA methylation is an attractive analyte for liquid biopsy because methylation patterns retain strong cell-lineage specificity.^1^ In plasma, cell free DNA (cfDNA) methylation has enabled cancer detection,^2^ tissue-of-origin inference^3^ and molecular subtyping.^4^ Yet practical translation remains constrained by several interrelated limitations.

First, tumor-derived cfDNA can occur at very low fractional abundance, particularly in low-burden disease,^5^ while hematopoietic cells contribute substantially to the circulating cfDNA background.^6^ Workflows that impose template loss or require extensive processing can therefore reduce analytical sensitivity precisely where detection is most challenging. For example, bisulfite-dependent methods can introduce additional processing and DNA damage,^7^ although recent methodological developments have shortened conversion workflows and improved recovery.^8^ Direct single-molecule detection of native modifications requires analysis of unamplified DNA molecules, making performance particularly dependent on recoverable template in low-input cfDNA settings.^9^

Second, informative methylation changes are often distributed across multiple loci,^10^ making the integration and interpretation of their combined signal an assay-design consideration. For low-input cfDNA applications, panel composition and measurement architecture must be considered together. Combining targets requires accounting for their individual methylation signals and background contributions, as well as defining how these contributions are represented in the final readout.^10,11^

Third, assay design must balance profiling breadth with workflow complexity. Broader methylation profiling approaches can capture distributed epigenetic information using enrichment-based cfDNA methylome profiling,^12^ targeted conversion-based methylation sequencing,^13^ enzymatic methylation sequencing^14,15^ and direct single-molecule analysis of native cfDNA.^9^ Bisulfite-free whole-genome approaches can preserve multimodal cfDNA information but remain sequencing-based.^16^ These broader profiling strategies can increase workflow and analytical complexity in routine implementation, with cost-effectiveness also depending on the sequencing effort required to obtain informative coverage.^17^

Here we developed Delta-HLD to quantify methylation directly in native DNA through sequential hybridization, ligation and methylation-sensitive digestion. In low-template settings such as plasma cfDNA, the primary objective is reliable detection of rare tumor-derived molecules against a large non-tumor background while accommodating biological heterogeneity across loci.⁵˒⁶ The assay couples locus-specific hemi-probe recognition to a shared amplification and detection architecture that co-reports multiple methylation targets as a single panel-level signal without base conversion. The context-aware framework informing candidate-region selection is described in the companion study by Angeloni et al.¹⁸, whereas the present study develops the architecture for their joint measurement.

## Results

### Delta-HLD converts native-DNA methylation into a panel-level PCR readout

Delta-HLD operates through three sequential steps: hybridization (H), ligation (L) and methylation-sensitive digestion (D) (Fig. 1a). For each target locus, two adjacent hemi-probes (HPs) recognize a native DNA template, with one hemi-probe spanning an HhaI-sensitive GCGC site. Correctly hybridized probe pairs are ligated to form an amplifiable product, which is then quantified across matched no-enzyme (NE) and enzyme-treated (E) reactions (Extended Data Fig. 1a). In the E condition, unmethylated molecules are cleaved and therefore lost from amplification, whereas methylated molecules remain intact and amplifiable. Because all Delta-HLD ligation products share a common primer/probe architecture, signal from multiple loci can be co-reported within a single fluorescence channel, and methylation is inferred from the relative signal in the paired reactions (Extended Data Fig. 1b). The primary output of a multilocus Delta-HLD configuration is therefore a co-reported methylation signal reflecting the aggregate methylation-dependent template retention across the interrogated tumor-associated loci.

**Figure 1.**
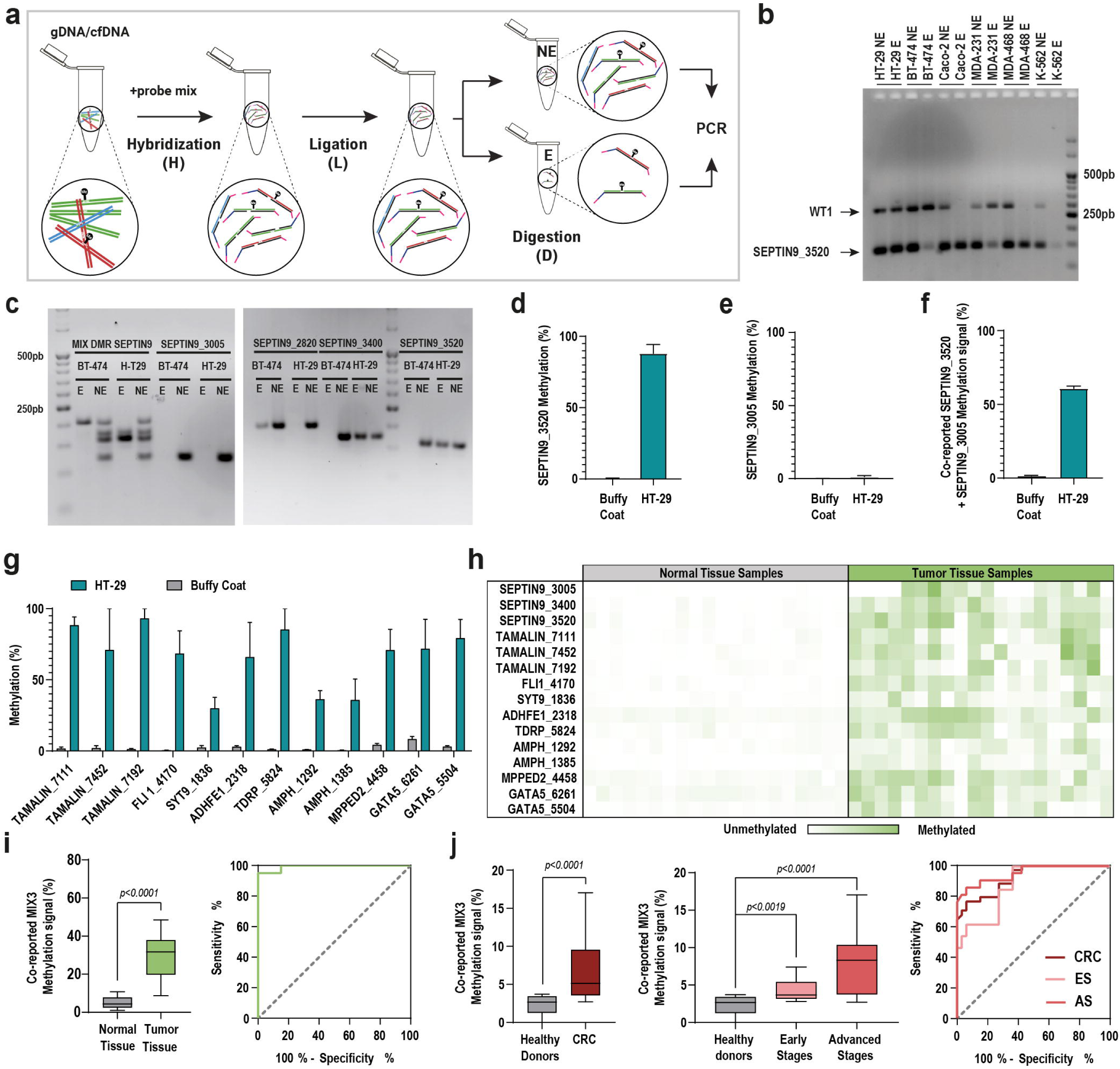
Delta-HLD chemistry, signal integration and preliminary CRC proof of concept. **a,** Schematic Delta-HLD workflow showing sequential hybridization, ligation and methylation-sensitive digestion with matched no-enzyme (NE) and enzyme-treated (E) reactions. **b,** Agarose gel analysis of SEPTIN9_3520 after Delta-HLD processing in DNA from six cancer cell lines, showing cell-line-dependent retention or suppression of amplification under no-enzyme (NE) and enzyme-treated (E) conditions, consistent with methylation-dependent protection from HhaI digestion. WT1 was included as a methylation-responsive comparator locus. **c,** Agarose gel analysis of individual SEPTIN9 Delta-HLD targets and a pooled SEPTIN9 configuration in BT-474 and HT-29 DNA under matched no-enzyme (NE) and enzyme-treated (E) conditions. The individual targets included SEPTIN9_3005, SEPTIN9_3400, SEPTIN9_3520 and SEPTIN9_2820 within the candidate methylation region prioritized in the accompanying discovery study. Target-specific length tags enabled size-based visualization of individual and pooled amplification products. **d-f,** Bar plots showing qPCR-based Delta-HLD methylation measurements for individual SEPTIN9_3520 (d), individual SEPTIN9_3005 (e), and the co-reported SEPTIN9_3520/3005 two-target configuration (f) in buffy coat DNA and HT-29 DNA. Individual-target methylation percentages and the two-target co-reported methylation signal were calculated from matched no-enzyme (NE) and enzyme-treated (E) reactions; bars show mean ± s.e.m. from technical replicates. **g,** Bar plots showing qPCR-based Delta-HLD methylation percentages for 12 CRC-associated hemi-probe systems in HT-29 DNA and human buffy coat DNA. Methylation percentages were calculated from matched no-enzyme (NE) and enzyme-treated (E) reactions; bars show mean ± s.e.m. from technical replicates. **h,** Heat map showing qPCR-based Delta-HLD validation of 15 targets comprising 12 CRC-associated loci and three SEPTIN9 targets in fresh-frozen colorectal cancer tumors and matched normal tissues (n = 20 pairs). Rows correspond to individual targets and columns to individual tissue samples; color intensity indicates methylation percentage. **i,** qPCR-based tissue-level validation of the empirically assembled three-locus configuration (SEPTIN9_3520, ADHFE1_2318 and FLI1_4170) in fresh-frozen matched colorectal cancer and normal colonic samples. Left, box plot showing the MIX3 co-reported methylation signal in matched normal and colorectal tumor tissues. The co-reported readout was significantly higher in tumors than in matched normal tissues (two-tailed exact Wilcoxon matched-pairs signed-rank test, P < 0.0001; n = 20 pairs). Right, ROC curve of tumor-normal tissue discrimination using the same co-reported MIX3 readout. **j,** Plasma performance of the same 3-locus MIX3 configuration during preliminary ddPCR-based assay development. Left, box plot showing the MIX3 co-reported methylation signal in healthy donors (n = 33) and patients with CRC (n = 34); groups were compared using a two-tailed Mann-Whitney test. Middle, box plot showing stage-stratified MIX3 co-reported methylation signal in healthy donors (n = 33), early-stage CRC (stages I-II; n = 13) and advanced-stage CRC (stages III-IV; n = 21); groups were compared using a Kruskal - Wallis test followed by Dunn’s multiple-comparisons test, with adjusted P values indicated in the panel. Right, ROC curves for overall CRC (n = 34), early-stage CRC (n = 13) and advanced-stage CRC (n = 21), each compared against healthy donors (n = 33).

We established the Delta-HLD chemistry using SEPTIN9, a canonical colorectal cancer (CRC) methylation target and a benchmark for blood-based methylation assays^19^. Using a Delta-HLD design centered on SEPTIN9_3520 (Chr 17: 77373520), PCR followed by agarose gel electrophoresis showed cell-line-dependent retention or suppression of amplification under enzyme-treated conditions, consistent with methylation-dependent protection from HhaI digestion (Fig. 1b). These results also showed that Delta-HLD can integrate methylation-dependent signals from multiple target sites into a single panel-level output. BT-474 and HT-29 cell lines were selected for the subsequent experiment because they showed distinct SEPTIN9_3520 Delta-HLD patterns, including an unmethylated profile in BT-474 and methylation-dependent protection and signal retention in HT-29. We then tested whether signal from multiple SEPTIN9 targets located within the SEPTIN9 candidate methylation region previously prioritized in the accompanying discovery study^18^ (Extended Data Fig. 1c) could be co-reported while preserving target- and cell-line-dependent signal differences. A set of SEPTIN9 hemi-probe systems spanning four positions within the prioritized region (Chr 17: 77373520; 77373400; 77373005; 77372820) showed that signals from several targets could be co-reported while preserving target-specific digestion-dependent signal behavior (Fig. 1c).

We next quantified locus-specific SEPTIN9_3520 methylation across six cancer cell lines using qPCR-based Delta-HLD. The resulting methylation profile was consistent with the cell-line-dependent signal pattern observed by gel electrophoresis (Extended Data Fig. 1d). We then quantified methylation at individual SEPTIN9 targets in HT-29 DNA and healthy-donor buffy coat-derived genomic DNA. SEPTIN9_3520 showed high methylation in HT-29 DNA and low signal in buffy coat DNA, whereas SEPTIN9_3005 showed low methylation in both DNA backgrounds (Fig. 1d,e). With these locus-specific quantitative readouts in place, we tested whether heterogeneous SEPTIN9 methylation states could be integrated into a single panel-level readout. When the high-methylation SEPTIN9_3520 target and the low-methylation SEPTIN9_3005 target were combined, the co-reported methylation signal shifted accordingly (Fig. 1f), supporting the ability of Delta-HLD to integrate heterogeneous site-specific methylation states into a single quantitative readout. Together, these experiments showed that the hybridization-ligation-digestion (HLD) sequence can convert native-DNA methylation states into methylation-dependent template retention after HhaI digestion, producing a PCR-detectable quantitative signal from matched NE and E reactions. The HLD architecture was also compatible with both qPCR and ddPCR (Extended Data Fig. 1e,f).

### Multi-locus co-reporting extends from tissue to plasma cfDNA

To extend Delta-HLD beyond a single benchmark gene, we tested whether candidate methylation targets selected during assay development could be operationalized in this assay format by leveraging a previously defined set of Delta-HLD hemi-probe systems targeting those loci, with candidate selection informed by the accompanying discovery study.^18^ We first verified their expected behavior in HT-29 DNA relative to buffy coat DNA (Fig. 1g), establishing assay-level compatibility in a tumor-derived DNA background and a low-methylation hematopoietic background. We then applied Delta-HLD to 15 targets, comprising 12 CRC-associated loci and three SEPTIN9 targets in fresh-frozen colorectal tumor tissues and matched normal-adjacent tissues (n = 20 pairs; Supplementary Table 1), detecting higher methylation signals in tumors than in matched normal-adjacent tissues (Fig. 1h).

To determine whether this information could be integrated into a panel-level readout, we next tested an empirically assembled simultaneous three-locus configuration, with targets located on three different chromosomes, comprising SEPTIN9_3520, ADHFE1_2318 and FLI1_4170 (MIX3). This tissue experiment was designed specifically to test multi-locus signal integration. The co-reported MIX3 readout retained strong tumor-normal discrimination in tissue, with an AUC of 0.9925 (95% CI, 0.9745-1.000; P < 0.0001), and was significantly higher in tumors than in matched normal-adjacent tissues (two-tailed Wilcoxon matched-pairs signed-rank test, P < 0.0001; n = 20 tissue pairs) (Fig. 1i).

We then asked whether the same MIX3 signal could be detected in plasma cfDNA using ddPCR, which provided direct signal visualization in the preliminary plasma workflow (Extended Data Fig. 1g). In plasma cfDNA, the MIX3 Delta-HLD signal discriminated CRC from healthy donors with an AUC of 0.9220 (95% CI 0.8624 - 0.9817; P < 0.0001; n = 33 healthy donors and 34 CRC cases; Supplementary Table 2). Stage-stratified analysis showed that discrimination was maintained in early-stage disease (AUC 0.8741, 95% CI 0.7700 - 0.9782; CRC = 13) and was higher in advanced-stage disease (AUC 0.9517, 95% CI 0.8955 - 1.000; CRC = 21) (Fig. 1j). Concordantly, this signal differed across groups (Kruskal-Wallis P < 0.0001) and was significantly higher than in healthy donors for both early-stage cases (Dunn’s adjusted P = 0.0019) and advanced-stage cases (Dunn’s adjusted P < 0.0001). Together, these data established plasma feasibility of the empirically assembled MIX3 configuration.

### Model-guided optimization defines a five-locus co-reported panel

We next aimed to develop a more transferable qPCR workflow. Before implementing the assay in this format, we first optimized the co-reported panel by defining its size and composition. To guide panel optimization, we used a model-based framework to determine the number and combination of target loci that could be co-reported while balancing discrimination against background. Candidate selection for model-based panel exploration was informed by the accompanying discovery study.^18^ Artificial tumor and normal methylation profiles were generated from GDC-TCGA β-values across 18 CpG loci. ^20,21^ To represent inter-tumor methylation heterogeneity, seven independently sampled tumor-profile sets were generated and analyzed in separate simulation runs. These profiles were then processed through a Repast/COPASI-based modeling framework that incorporated assay-related uncertainty terms, including DNA recovery, dilution, incomplete enzymatic digestion and qPCR specificity (Fig. 2a; representative methylation distributions for CpG sites 1 and 10 across the seven independently sampled tumor-profile sets are shown in Extended Data Fig. 2a; Methods). Across seven independent simulations, multi-locus panels consistently outperformed single-locus assays, but gains plateaued with increasing panel size, identifying an optimal range of 5 - 8 loci (Fig. 2b and Extended Data Fig. 2b). To translate this into an experimentally implementable assay, we restricted the analysis to the 15 loci compatible with the Delta-HLD (Fig. 1h), as three candidates lacked a nearby GCGC site required for HhaI-sensitive target selection. Using Delta-HLD methylation values obtained from the 20 matched CRC tumor-normal tissue pairs shown in Fig. 1h, we ranked all possible 8-locus combinations according to their tumor-normal AUC and identified recurrent candidate panels for experimental testing (Fig. 2c).

**Figure 2.**
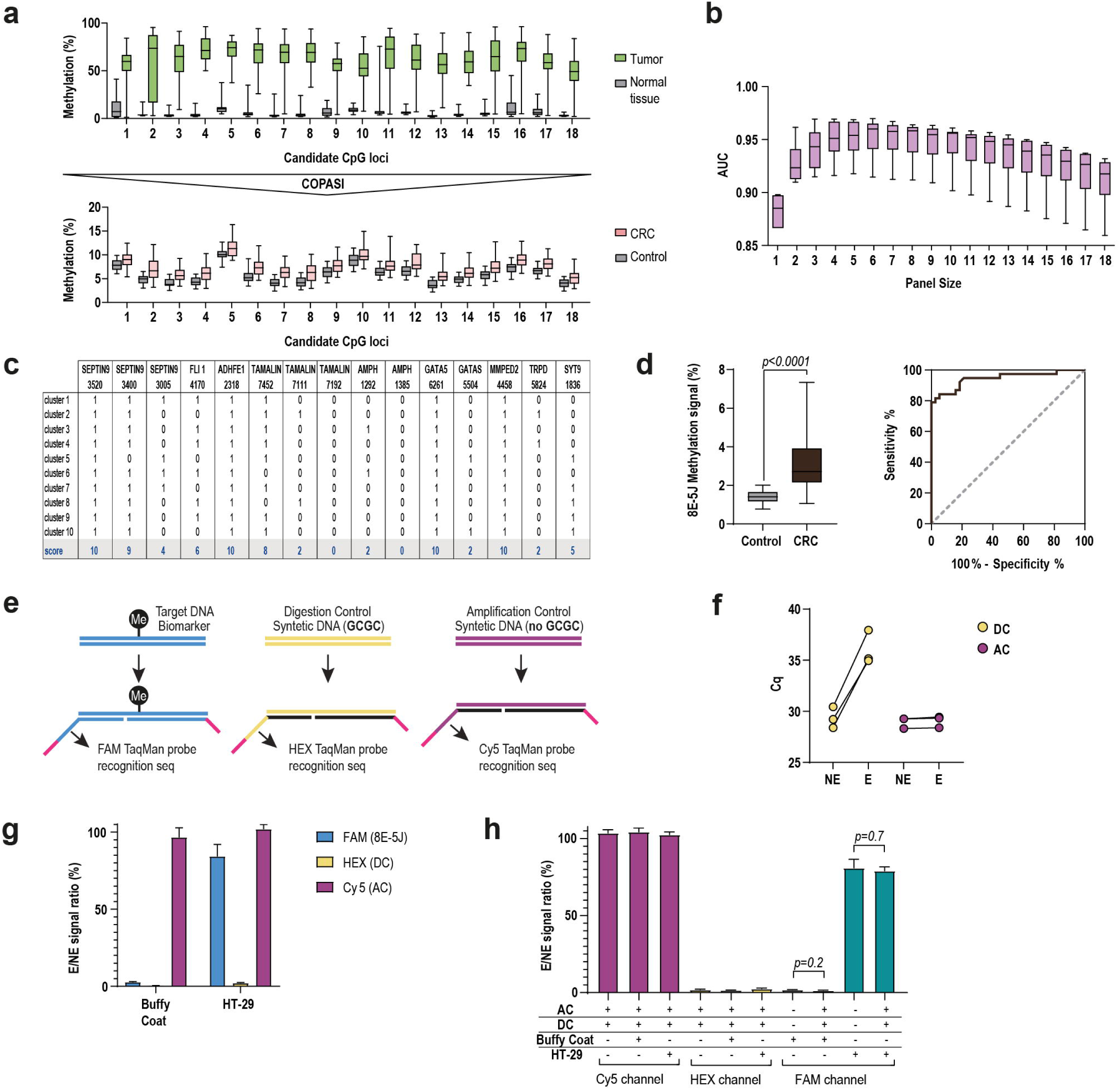
Model-guided optimization of panel composition and implementation of the final triplex qPCR workflow. **a,** Box plots showing simulated methylation profiles across 18 CpG loci used to model the effect of panel size and composition under assay-related uncertainty terms. Top, tumor and normal tissue methylation profiles used as model input. Bottom, corresponding CRC and control profiles after COPASI-based simulation under assay-related uncertainty terms. **b,** Box plots summarizing modeled classification performance across seven independent simulations, showing that multi-locus panels outperform single-locus assays and plateau within an optimal range of 5 - 8 co-reported loci. **c,** Binary inclusion matrix showing recurrent candidate 8-locus configurations identified from the assay-compatible subset of targets. Values indicate whether each locus was included in each candidate configuration; the bottom row summarizes recurrence across configurations. **d,** Performance of the optimized 8E-5J co-reported configuration in expanded CRC-derived artificial liquid biopsies. Left, box plot showing co-reported methylation signal in matched control-spiked and tumor-spiked artificial liquid biopsies. Groups were compared using a two-tailed exact Wilcoxon matched-pairs signed-rank test (P < 0.0001; n = 38 pairs). Right, ROC curve for tumor-spiked sample discrimination, n = 38 control-spiked and n = 38 tumor-spiked samples. **e,** Schematic of the final triplex qPCR architecture, in which the 8E-5J target panel is measured in the FAM channel, a synthetic digestion control (DC) containing an HhaI site is measured in HEX and a synthetic amplification control (AC) lacking an HhaI site is measured in Cy5. **f,** Paired dot plot showing DC and AC Cq values in matched no-enzyme (NE) and enzyme-treated (E) reactions. Individual Cq values are shown for three technical replicates per condition, with connecting lines indicating matched NE/E reaction pairs. DC showed the expected digestion-dependent Cq shift after enzyme treatment, whereas AC remained comparable between NE and E reactions. **g,** Bar plots showing complete triplex behavior in buffy coat and HT-29 DNA, shown as E/NE signal ratio for the 8E-5J target-panel channel (FAM), digestion-control channel (HEX) and amplification-control channel (Cy5). Bars show mean + s.e.m. from three technical replicates. Low FAM signal in buffy coat, high FAM signal in HT-29, suppressed HEX signal and preserved Cy5 signal support the expected behavior of the three-channel system. **h,** Bar plots showing compatibility of DC and AC incorporation with the FAM panel readout. The plus and minus symbols indicate the presence or absence of AC, DC, buffy coat DNA and HT-29 DNA in each reaction condition. E/NE signal ratios are shown for Cy5, HEX and FAM channels. Bars show mean + s.e.m. from three technical replicates. Addition of DC and AC preserved the expected FAM panel readout in buffy coat DNA and HT-29 DNA, while AC retained the expected high E/NE signal ratio in Cy5 and DC remained strongly suppressed in HEX.

We next tested these model-guided predictions in artificial liquid biopsies generated by spiking either CRC tumor or matched non-tumor tissue DNA into a constant buffy coat DNA background. Ten 8-locus co-reported panels (8A to 8J) were initially screened by ddPCR in no-template reactions to quantify baseline HP-system noise, and the two lowest-background configurations, 8A and 8E, were selected for further evaluation in DNA-containing samples (Extended Data Fig. 2c,d). Although both 8A and 8E discriminated tumor-spiked from control-spiked samples, broader co-reporting increased baseline signal in controls, indicating excess background under the 8-locus format (Extended Data Fig. 2e). Guided by the model, we therefore reduced panel size to 5 loci and tested four 5-locus derivatives of the selected 8-locus panels: 8A-5C, 8A-5E, 8E-5A and 8E-5J. This lowered baseline signal in controls and improved tumor-control separation. The 8E-5J configuration, comprising SEPTIN9_3520, FLI1_4170, TAMALIN_7452, GATA5_6261 and MPPED2_4458, was selected for subsequent assay development (Extended Data Fig. 2f). The selected configuration was subsequently evaluated in an expanded set of CRC-derived artificial liquid biopsies generated from 38 matched tumor–normal tissue pairs (Supplementary Table 3), where it retained low control background and achieved robust discrimination (AUC = 0.9456, 95% CI 0.8932–0.9981; Fig. 2d). Under the same qPCR-based artificial liquid-biopsy conditions used for the model-guided panel screening, the original three-locus MIX3 configuration also discriminated tumor-spiked from control-spiked samples, but generated a lower co-reported methylation signal than the 8E-5J configuration (Extended Data Fig. 2g). Direct head-to-head comparison of MIX3 and 8E-5J showed a higher co-reported methylation signal with 8E-5J (Extended Data Fig. 2h), and this increase was consistent across eight independent tumor-derived artificial liquid-biopsy samples analyzed in parallel (Extended Data Fig. 2i).

### Triplex qPCR combines panel detection with internal process controls

Having defined 8E-5J as the optimized co-reported configuration, we next focused on transferring this panel into a qPCR format compatible with routine laboratory workflows. Because Delta-HLD methylation is inferred from matched NE/E reactions, this transition required internal controls capable of distinguishing true methylation-dependent template retention from altered digestion efficiency or amplification performance. We therefore implemented a triplex qPCR configuration, where the 8E-5J target panel is measured in the FAM channel, a digestion control (DC) in HEX and an amplification control (AC) in Cy5 (Fig. 2e).

DC consists of a synthetic template containing a single HhaI site; effective digestion is indicated by marked depletion of the E/NE signal ratio. By contrast, AC lacks an HhaI site and should maintain comparable amplification between NE and E, thereby controlling for reaction integrity and amplification comparability across the matched pair. We first optimized the DC template for the qPCR implementation, selecting 0.015 pg/µl as an optimal concentration that ensured reliable detection while minimizing potential competition with the panel reaction (Extended Data Fig. 3a). A parallel ddPCR titration confirmed the expected digital behavior of the same control (Extended Data Fig. 3b). Representative ddPCR amplitude plots and concentration estimates verified channel-specific detection of the 8E-5J target panel and DC in the FAM and HEX channels (Extended Data Fig. 3c,d). We then implemented the complete triplex qPCR configuration by incorporating AC as a third channel, with the 8E-5J target measured in FAM, DC in HEX and AC in Cy5. In this format, DC showed a marked Cq shift between NE and E reactions, whereas AC remained comparable across the matched pair, consistent with their intended control functions (Fig. 2f). Representative qPCR amplification profiles further confirmed triplex detection of the 8E-5J target panel, DC and AC in the FAM, HEX and Cy5 channels, respectively (Extended Data Fig. 3e).

In the complete triplex system, buffy coat DNA showed minimal FAM signal, strongly suppressed HEX signal and preserved high Cy5 signal, whereas HT-29 DNA showed the expected 8E-5J target panel FAM signal together with similarly suppressed HEX signal and preserved high Cy5 signal (Fig. 2g). Finally, incorporation of both DC and AC was compatible with the FAM panel readout, as the FAM E/NE signal ratio remained comparable in buffy coat DNA and HT-29 DNA under the complete triplex configuration (Fig. 2h). Under the same conditions, AC retained the expected high E/NE signal ratio in the Cy5 channel and DC remained strongly suppressed in HEX, supporting the expected performance of both control channels in the presence of sample DNA.

### Optimized Delta-HLD qPCR generates informative plasma signals in CRC and HCC

With the model-guided 8E-5J configuration and triplex qPCR architecture defined, we next adapted Delta-HLD for plasma cfDNA analysis. This optimization combined preanalytical changes, including stabilized blood collection, increased plasma input and magnetic bead-based cfDNA extraction, with an analytical input-range evaluation obtained from the final triplex qPCR workflow (Extended Data Fig. 4a-c). Across serial DNA inputs, FAM NE Cq values increased as input declined, whereas HEX and Cy5 control behavior remained stable, supporting use of Cq(NE) as a sample-level input criterion. Spike-in experiments in a fixed unmethylated background further defined a low-input detectability range for methylated control DNA. Based on these studies, an input-acceptance criterion was established for the final triplex qPCR workflow (Extended Data Fig. 4d-f) and incorporated into the optimized plasma analysis workflow (Fig. 3a). Samples outside the defined input-acceptance range were excluded from downstream plasma analyses.

**Figure 3.**
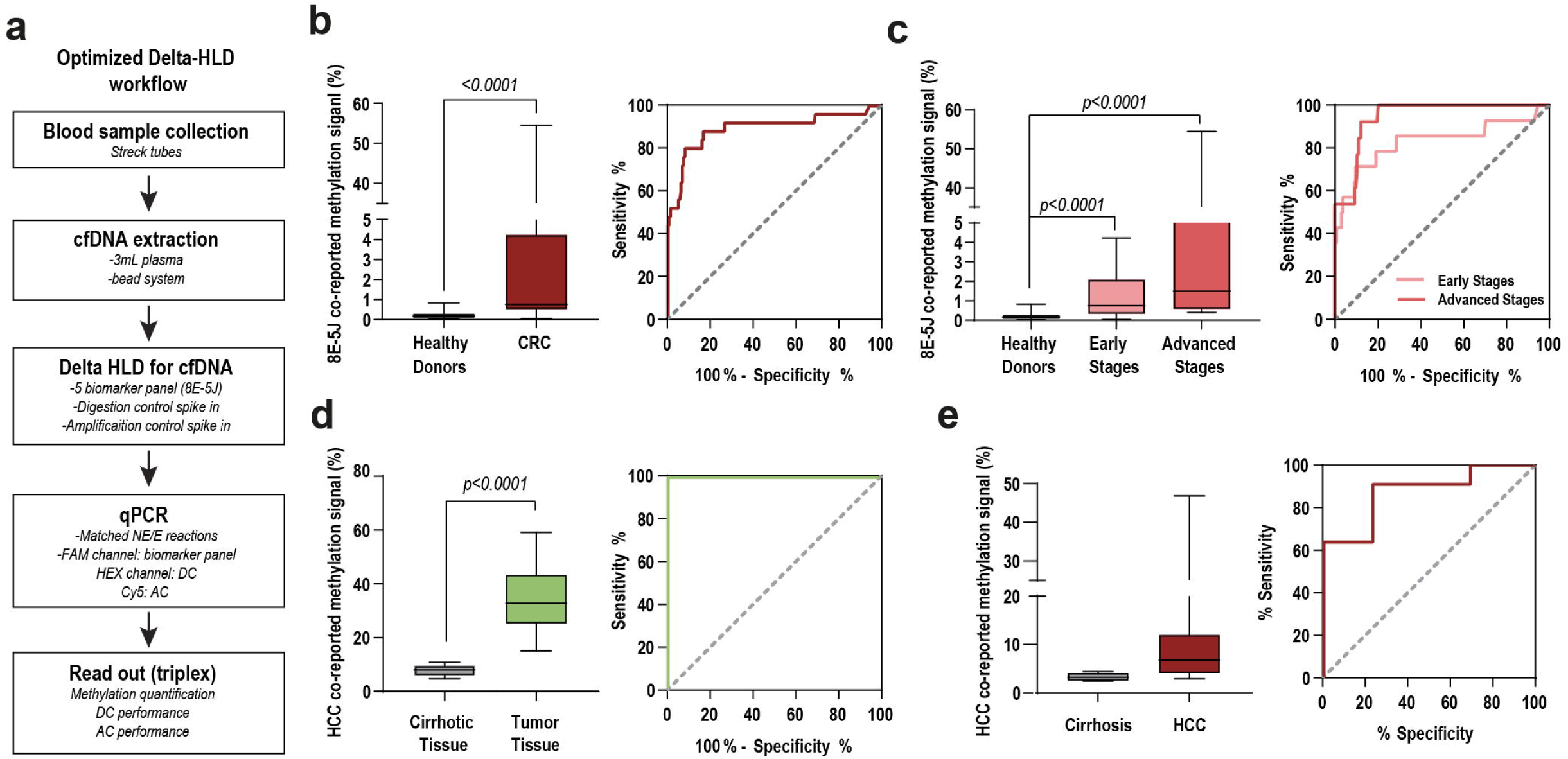
Plasma proof-of-concept of the optimized qPCR Delta-HLD workflow and application to HCC. **a,** Schematic of the optimized Delta-HLD workflow for plasma cfDNA analysis, including Streck blood-collection tubes, magnetic bead-based cfDNA extraction from 3 ml plasma, the 5-locus 8E-5J Delta-HLD panel with digestion-control (DC) and amplification-control (AC) spike-ins, matched no-enzyme (NE) and enzyme-treated (E) qPCR reactions and triplex readout with per-sample input acceptance based on FAM Cq (NE). **b,** Plasma performance of the optimized 5-locus 8E-5J CRC configuration in a CRC plasma proof-of-concept sample set. Left, box plot showing co-reported 8E-5J methylation signal in healthy donors (n = 164) and patients with CRC (n = 25). Right, ROC curve for CRC discrimination from healthy donors. **c,** Stage-stratified CRC plasma analysis. Left, box plot showing co-reported 8E-5J methylation signal in healthy donors (n = 164), early-stage CRC (n = 13) and advanced-stage CRC (n = 12). Right, ROC curves for early-stage and advanced-stage CRC, each compared with healthy donors. **d,** Co-reported methylation signal and ROC analysis for the HCC panel in cirrhotic liver tissue (n = 12) and HCC tumor tissue (n = 11). **e,** Co-reported methylation signal and ROC analysis in plasma cfDNA from patients with cirrhosis (n = 13) and HCC (n = 11).

We next asked whether the optimized 5-locus 8E-5J configuration retained informative signal in a plasma sample set comprising patients with CRC and healthy donors. Delta-HLD was evaluated in the triplex qPCR format using plasma cfDNA from patients with CRC (n = 25) and healthy donors (n = 164; Supplementary Table 4). Delta-HLD discriminated CRC from healthy donors with an AUC of 0.8985 (95% CI, 0.8111-0.9860; P < 0.0001; Fig. 3b). Stage-stratified analysis showed that the 8E-5J co-reported methylation signal differed across healthy donors, early-stage CRC and advanced-stage CRC. Both early-stage CRC and advanced-stage CRC showed significantly higher co-reported methylation signals than healthy donors (two-tailed exact Mann-Whitney tests, P < 0.0001 for both comparisons), with AUCs of 0.8422 for early-stage CRC (95% CI, 0.6845-0.9999; P < 0.0001; n = 13 vs 164 healthy donors) and 0.9596 for advanced-stage CRC (95% CI, 0.9259-0.9933; P < 0.0001; n = 12 vs 164 healthy donors) (Fig. 3c).

We then asked whether the same platform logic could be transferred beyond CRC. As a proof of concept in a distinct tumor context, we applied qPCR Delta-HLD to hepatocellular carcinoma (HCC), a biologically distinct setting using cirrhosis as a disease-relevant non-malignant comparator^22^. Assay-compatible HCC targets selected for analytical evaluation showed tumor-associated methylation enrichment relative to cirrhotic liver tissue samples (Extended Data Fig. 5). We then evaluated a co-reported 5-locus HCC configuration comprising IDUA_2443, USP44_8196, USP44_8324, CPAMD8_7837 and CPAMD8_7923. In tissue, the panel showed clear tumor-cirrhosis separation with an AUC of 1 (n = 11 HCC tumor tissues and n = 12 cirrhotic liver tissues; Supplementary Table 5; Fig. 3d). The same HCC configuration was subsequently tested in an HCC plasma proof-of-concept sample set and distinguished patients with HCC from patients with cirrhosis, achieving an AUC of 0.8741 (95% CI 0.7274 - 1.000; P = 0.0019; 11 HCC versus 13 cirrhotic patients; Supplementary Table 6; Fig. 3e). Together, these analyses show that the optimized Delta-HLD qPCR implementation can generate informative plasma signal in a second tumor context without changing the underlying assay logic.

### Delta-HLD optimization for cfDNA analysis following platelet-retaining processing

During assembly of the plasma sample sets used to evaluate qPCR-based Delta-HLD in plasma cfDNA, Murphy et al. reported that platelet-depleted plasma can discard a potentially informative fraction of extracellular DNA^23^. This observation led us to hypothesize that Delta-HLD signal recovery might be improved by increasing recovery of platelet-associated circulating DNA. We therefore tested a platelet-retaining sample-processing protocol that differed from our standard cfDNA extraction workflow primarily at the centrifugation step (Fig. 4a). Macroscopic inspection showed that the standard protocol produced a clear plasma fraction with a sharply defined buffy coat layer, whereas the platelet-retaining protocol yielded a more turbid plasma fraction and a less distinct buffy coat, suggesting reduced platelet depletion but also a higher risk of genomic DNA carryover. DNA extracted from the resulting plasma fractions showed markedly higher concentrations under the platelet-retaining protocol (Fig. 4b), consistent with increased total DNA recovery.

**Figure 4.**
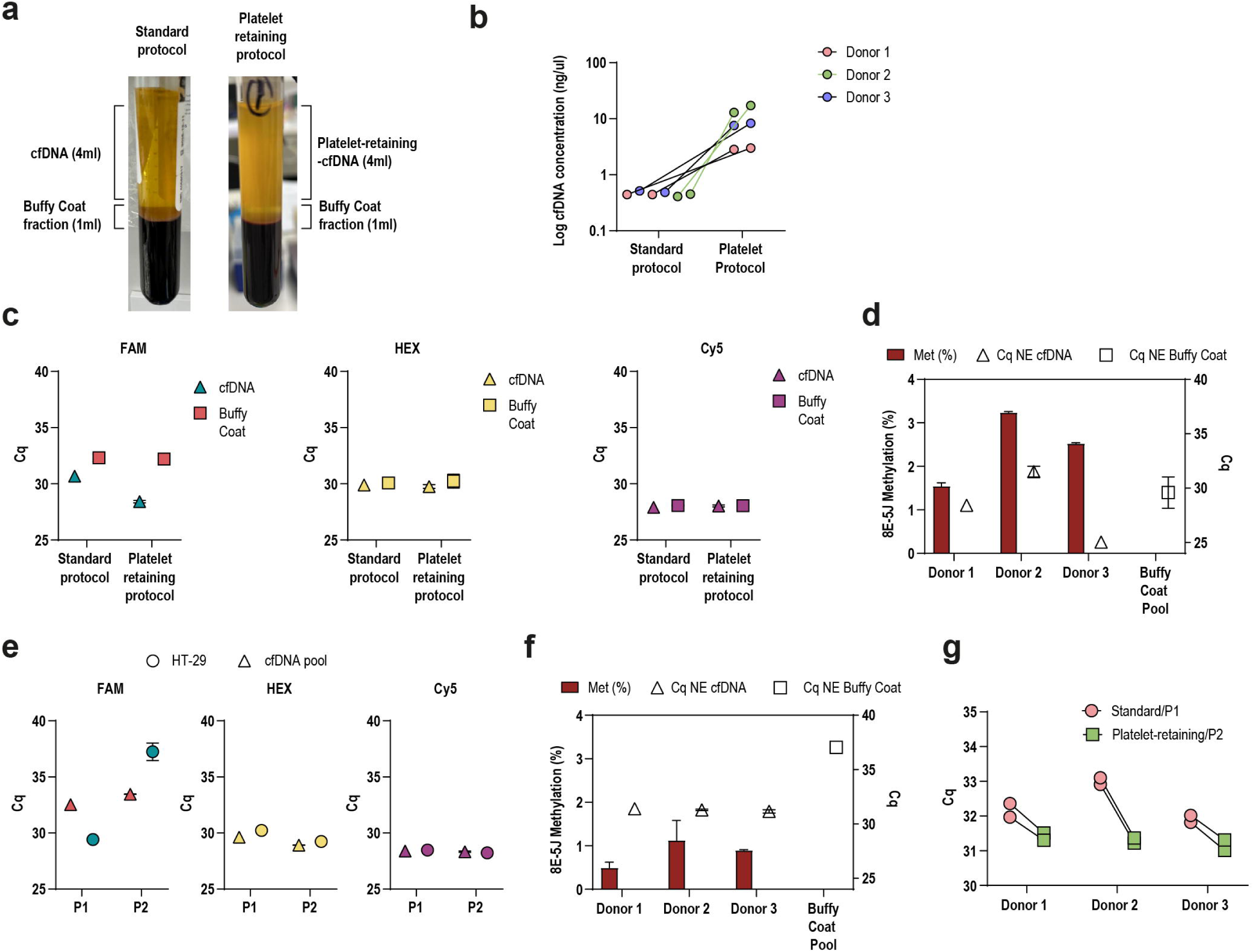
Platelet-retaining processing and P2 refinement improve Delta-HLD recovery of circulating templates. **a,** Representative images comparing the standard cfDNA processing protocol and the platelet-retaining protocol, designed to increase recovery of platelet-associated circulating DNA before Delta-HLD analysis. The standard protocol produced a clear plasma fraction and a sharply defined buffy coat layer, whereas the platelet-retaining protocol yielded a more turbid plasma fraction and a less distinct buffy coat layer. **b,** Paired dot plot showing total DNA concentration recovered from circulating fractions from three healthy donors processed with the standard and platelet-retaining protocols. Lines connect paired donor samples. **c,** Dot plots showing Cq values from qPCR-based Delta-HLD analysis of circulating DNA and buffy coat-derived DNA fractions obtained with the standard and platelet-retaining protocols using the original P1 protocol. Cq values are shown for the no-enzyme (NE) reaction in the methylation-target channel (FAM), digestion-control channel (HEX) and amplification-control channel (Cy5). The platelet-retaining protocol produced earlier FAM NE Cq values in the circulating DNA fraction while preserving control-channel behavior. **d,** Combined bar and symbol plot showing initial Delta-HLD analysis of three healthy-donor platelet-retaining cfDNA samples and a pooled buffy coat DNA control under the platelet-retaining protocol using P1. Bars show the 8E-5J co-reported methylation signal; symbols show FAM NE Cq values for cfDNA and buffy coat DNA. **e,** Dot plots showing Cq values from qPCR-based Delta-HLD analysis comparing the original P1 protocol and the selected P2 protocol using genomic HT-29 DNA and pooled healthy-donor cfDNA. Genomic HT-29 DNA was used as a model of high-molecular-weight DNA carryover, whereas pooled cfDNA was used as a model of short circulating templates. Cq values are shown for FAM, HEX and Cy5 channels, demonstrating reduced genomic HT-29 DNA detection under P2 while preserving cfDNA-associated signal and control-channel behavior. **f,** Combined bar and symbol plot showing reanalysis of three healthy-donor platelet-retaining cfDNA samples and pooled buffy coat DNA under the selected P2 protocol. Bars show 8E-5J co-reported methylation signal; symbols show FAM NE Cq values for cfDNA and buffy coat-derived genomic DNA background illustrating reduced apparent methylation and reduced recognition of buffy coat-derived genomic DNA background. **g,** Paired dot plot comparing healthy-donor samples processed under two complete conditions, standard cfDNA extraction followed by the original P1 Delta-HLD protocol, and platelet-retaining extraction followed by the selected P2 protocol. FAM NE Cq values are shown for the circulating DNA fraction from each donor; lines connect paired donor samples, showing earlier qPCR signal under the platelet-retaining P2 condition.

When these fractions were analyzed by qPCR Delta-HLD, the circulating fraction obtained with the platelet-retaining protocol showed a clear gain in analyzable template, reflected by earlier FAM NE Cq values, whereas buffy coat DNA remained comparatively stable (Fig. 4c). At the same time, HEX and Cy5 behavior remained stable, indicating that the internal-control architecture was preserved despite the pre-analytical change. However, when this workflow was extended to three healthy donors, the resulting 8E-5J co-reported methylation signals were unexpectedly elevated in healthy controls and donor-to-donor FAM NE Cq values became highly variable, whereas a pooled buffy coat DNA control behaved differently (Fig. 4d). These findings suggested that the gain in total recovered DNA was accompanied by increased genomic DNA contamination, which could inflate apparent methylation signals and compromise biological interpretability.

On the basis of this observation, we next asked whether the early steps of Delta-HLD could be modified to increase specificity for short circulating fragments while reducing genomic DNA recognition. We compared three reaction schemes, comprising the original P1 protocol and two modified protocols, P2 and P3, that altered the initial thermal-processing steps before the common hybridization, ligation and digestion sequence (Extended Data Fig. 6a).

P2 eliminated the initial 5-min high-temperature DNA denaturation and the separate 25 °C incubation after probe addition, whereas P3 retained the P1 sequence but reduced both high-temperature steps to 80 °C. Genomic HT-29 DNA was used as a model of high-molecular-weight DNA carryover, and short synthetic DC/AC templates were used as models of short fragments. Under these conditions, P2 and P3 reduced genomic DNA detection relative to the original P1 condition while maintaining control detection. We selected P2 because it simplified the workflow while preserving robust detection of short control templates (Extended Data Fig. 6b).

We next evaluated P1 and P2 using pooled cfDNA from healthy donors as a source of short circulating templates and genomic HT-29 DNA as a model of non-informative high-molecular-weight DNA carryover. The selected P2 protocol preserved cfDNA-associated FAM signal while reducing genomic HT-29 DNA detection, with stable HEX and Cy5 control behavior (Fig. 4e). Reanalysis of healthy-donor samples under P2 showed a marked reduction in the apparent 8E-5J co-reported methylation signal relative to the initial condition, reaching values closer to those observed under the standard cfDNA workflow, whereas the buffy coat DNA pool no longer met the previously defined NE input criterion, consistent with reduced recognition of non-informative genomic DNA background (Fig. 4f). Finally, we compared the two complete processing-and-assay conditions in paired healthy-donor samples: standard cfDNA extraction followed by the original P1 Delta-HLD protocol, and platelet-retaining extraction followed by the P2 protocol. The platelet-retaining P2 condition produced earlier FAM NE Cq values in the circulating DNA fraction across donors (Fig. 4g), consistent with increased recovery of circulating DNA signal after assay re-optimization to reduce genomic DNA recognition.

Together, these findings support combining platelet-retaining processing with the P2 protocol to increase recoverable circulating DNA signal while reducing genomic DNA recognition.

## Discussion

When molecular information is distributed across multiple loci, no single site carries the entire message. Delta-HLD was designed to integrate these distributed methylation signals into a common panel-level readout while preserving native DNA for PCR-compatible analysis. DNA methylation can encode tumor-associated, tissue-of-origin and subtype-related information in cfDNA,^3,4^ and low circulating tumor fraction remains a fundamental constraint on detection in low-burden settings^5^. Through sequential hybridization, ligation and digestion, Delta-HLD converts methylation-dependent template retention into a quantitative readout from matched no-enzyme and enzyme-treated reactions. A shared amplification and detection architecture enables multiple tumor-associated targets to be co-reported, producing a co-reported methylation signal that integrates heterogeneous locus-specific inputs. This architecture is intended for low-input settings in which sensitive detection of low-abundance tumor-derived molecules is the primary analytical objective.

Previous targeted methylation assays have demonstrated complementary elements of this analytical space, including measurement of multiple methylation targets within a common fluorescence readout,^11,24^ locus-specific probe recognition coupled to common-primer amplification with individual products resolved by capillary electrophoresis,^25^ and direct or bisulfite-free PCR-based detection of methylation.^26,27^ Bisulfite-free multiplex methylation detection has also been implemented using a universal primer combined with target-specific terminal primers^28^. In Delta-HLD, locus-specific target recognition occurs at the hemi-probe stage, whereas methylation-dependent selection is mediated by HhaI digestion. Ligated products share a common amplification and detection architecture that intentionally co-reports their methylation-dependent contributions. The resulting quantitative signal integrates methylation-dependent retention across the selected targets within a single panel-level readout.

This shared amplification architecture also addresses the challenge of coordinating multiple locus-specific PCR systems within a single assay. Designing short amplicons in GC-rich methylation targets and coordinating multiple locus-specific primer pairs can constrain cfDNA assay development^29,30^. By placing locus-specific recognition in adjacent hemi-probes, Delta-HLD allows target panels to be reconfigured while retaining common PCR primers and a shared detection probe. This architecture avoids a separate PCR primer/probe design for each target, although local sequence requirements and the performance of individual hemi-probe systems and their combinations still require experimental evaluation.

Delta-HLD was implemented with both real-time PCR and digital PCR. ddPCR provides a partition-based framework for quantitative nucleic acid analysis and has also been applied to DNA methylation measurements^31,32^. We validated a duplex ddPCR implementation in which FAM reported the methylation-target panel and HEX monitored restriction digestion. We then implemented the optimized assay as a triplex qPCR workflow, with the target panel measured in FAM, a digestion control in HEX and an amplification control in Cy5. In this configuration, fluorescence channels support a single panel-level methylation readout together with orthogonal monitoring of digestion and amplification within a PCR format already embedded in translational laboratories. Importantly, the control logic is not intrinsically restricted to qPCR; multiplexed ddPCR implementations could incorporate additional orthogonal control or target channels in future assay configurations.

Panel composition was guided by model-based analysis followed by artificial liquid-biopsy experiments to balance heterogeneity capture against co-reporting-associated background, supporting rational selection of the optimized 5-locus 8E-5J configuration. Multilocus methylation assays have independently demonstrated the value of integrating distributed tumor-associated methylation signals in CRC.^10^ Our experiments further showed that increasing the number of co-reported targets does not necessarily improve performance when background rises in parallel. Thus, the final Delta-HLD workflow couples methylation-dependent template retention with rational panel-level signal integration and per-sample monitoring of digestion and amplification.

The CRC and HCC analyses provide patient-derived proof-of-concept evidence for this architecture. In CRC, Delta-HLD progressed from chemistry establishment and ddPCR-based plasma feasibility to an optimized triplex qPCR workflow using the 5-locus 8E-5J configuration. In the broader CRC field, prospective blood-based cfDNA testing has demonstrated CRC detection in a screening population^33^, and independent multicenter studies have supported targeted plasma methylation measurements for CRC detection.^34^ In HCC, the same assay logic was applied to a distinct tumor context and evaluated against cirrhosis, a disease-relevant non-malignant comparator. In this disease context, plasma methylation approaches have shown discrimination of HCC from non-malignant liver disease backgrounds^35^, and circulating methylated SEPTIN9 has recently been evaluated for HCC detection in patients with cirrhosis^22^. Within this broader context, the results presented here indicate that the Delta-HLD architecture can accommodate distinct target panels and tumor contexts while preserving the same underlying assay logic. These analyses remain proof-of-concept and do not constitute indication-specific clinical validation.

During the plasma proof-of-concept evaluation of Delta-HLD, platelets were reported to actively sequester extracellular tumor-derived DNA, making the recoverable cfDNA fraction dependent in part on preanalytical handling^23^. Independent methylation analyses have also identified lineage-specific cfDNA patterns in plasma and platelets arising from megakaryocyte and erythroblast biology^36^. These findings provided an opportunity to evaluate Delta-HLD under platelet-retaining conditions. This evaluation motivated refinement of the initial thermal-processing steps with the aim of favoring hemi-probe hybridization to short DNA templates while reducing genomic DNA recognition. In our experiments, the Delta-HLD P2 refinement reduced genomic DNA recognition while preserving cfDNA-associated signal and control-channel behavior. When coupled to platelet-retaining processing, P2 produced earlier FAM NE Cq values in circulating DNA than standard processing with the original protocol. This comparison evaluated the combined processing-and-assay condition. P2 represents an assay optimization, and its combination with platelet-retaining processing has not yet been clinically validated. These findings support evaluating DNA recovery together with assay recognition of circulating and genomic templates.

Several limitations should be considered. The CRC plasma analysis remains case-control and requires prospective evaluation with prespecified thresholds and locked workflows. The HCC cohort evaluated the assay architecture in a second tumor context and was not powered for clinical validation. The platelet-retaining and P2 experiments remain exploratory and do not yet demonstrate improved clinical sensitivity in patient samples. The CRC and HCC configurations evaluated here were derived from candidate spaces informed by public methylome cohorts and were assessed in local tissue cohorts, so their generalizability beyond these populations remains to be established^18^. The co-reported plasma panels evaluated here nevertheless require validation in independent and population-representative cohorts. This requirement is consistent with proposed translational frameworks for methylation biomarker development^37^. Delta-HLD is a compact native-DNA methylation-detection platform designed to integrate signals into a panel-level output. Indication-specific clinical validation will require dedicated prospective studies.

## Methods

### Study design, participants and ethics

Biological specimens were obtained from participants enrolled in two independent case-control studies conducted in Mendoza, Argentina: CASCADE for colorectal cancer (CRC; ClinicalTrials.gov identifier NCT07148297) and HIDE for hepatocellular carcinoma (HCC; NCT07148310). All participants provided written informed consent. Both studies were conducted in accordance with the Declaration of Helsinki and institutional regulations, with approval from the ethics committees of Hospital Español de Mendoza (Act No. 38/2022) and Hospital Central de Mendoza (Act No. 01/2023), respectively.

CASCADE participants were classified using colonoscopic evaluation, clinical-record review and histopathological confirmation, as applicable. Specimens from 256 participants were included across the experimental cohorts, comprising 59 CRC cases and 197 non-cancer controls. HIDE included 37 participants, comprising 22 with HCC and 15 with cirrhosis. Clinical and histopathological evaluation established the corresponding diagnoses, and cirrhosis served as the disease-relevant non-malignant comparator for HCC. Liver disease etiology was classified using the updated Steatotic Liver Disease nomenclature³⁸. Tissue and plasma specimens were analyzed in experiment-specific cohorts, with sample sizes and characteristics reported in the corresponding Results, figure legends and Supplementary Tables.

### Biological specimens and reference materials

Clinical materials included peripheral blood, fresh-frozen colorectal tumor and matched non-neoplastic mucosa, and fresh-frozen or archival formalin-fixed paraffin-embedded liver specimens. Tissue diagnoses were confirmed by pathologists, and paired tumor and non-malignant specimens were analyzed when available. Tissue subsets used for target-level evaluation overlapped with those examined in the accompanying discovery study¹⁸.

Cancer-cell-line DNA, pooled healthy-donor buffy-coat DNA and commercially available methylated and unmethylated reference DNA were used for assay development and analytical assessments. Reference materials were selected according to the methylation targets and experimental objectives.

### Sample processing and DNA preparation

Blood-processing and DNA-extraction workflows were evaluated during assay development, considering their effects on cfDNA recovery, composition and fragmentation³⁹,⁴⁰. Plasma was prepared using either standard platelet-depleted or platelet-retaining workflows, according to the experimental stage. Buffy-coat fractions were collected separately as a source of hematopoietic reference DNA.

DNA was isolated using commercial extraction systems appropriate for plasma, cultured cells, buffy coat, frozen tissue or archival tissue. Artificial liquid-biopsy samples were prepared by diluting DNA from CRC tumors or matched non-neoplastic mucosa into pooled buffy-coat DNA. These controlled mixtures supported experimental evaluation of the assay under low-abundance tissue-derived DNA conditions.

### Delta-HLD assay and PCR readout

Delta-HLD was performed on native genomic DNA or cfDNA without cytosine conversion. Locus-specific hemi-probes hybridized to adjacent regions of the DNA template and underwent target-dependent ligation. The resulting products shared a common amplification architecture, enabling individual targets or multiple loci to be quantified using a common PCR readout.

Following hybridization and ligation, reactions were divided into matched no-enzyme and enzyme-treated aliquots. Methylation-sensitive processing with HhaI selectively reduced products associated with unmethylated targets. Signals from the paired conditions were subsequently measured by droplet digital PCR (ddPCR) or quantitative PCR (qPCR).

Single-target measurements were expressed as methylation percentages. Multilocus measurements were reported as co-reported methylation signals reflecting aggregate methylation-dependent retention across the interrogated targets. The no-enzyme condition provided the corresponding total amplifiable target signal.

Synthetic digestion and amplification controls were incorporated during assay optimization to monitor processing and amplification performance. Initial experiments used single-target or co-reported target detection, whereas the optimized qPCR format simultaneously measured the methylation-target signal and both internal controls. Methylated, unmethylated and no-template controls were included, and qPCR samples were analyzed in at least duplicate reactions. Assay reporting followed digital MIQE and MIQE recommendations where applicable⁴¹,⁴².

### Analytical assessment and workflow optimization

Analytical experiments examined assay behavior across DNA inputs, methylation backgrounds and fragment characteristics. Alternative thermal-processing schemes were compared for their responses to short DNA templates and genomic DNA. The selected workflow for subsequent platelet-retaining plasma experiments preserved short-template detection while reducing genomic-DNA recognition.

Plasma samples were assessed for sufficient amplifiable input using the no-enzyme target readout. Input-limited samples were excluded from downstream analyses. Internal controls were used to assess digestion and amplification performance.

TCGA-derived methylation profiles were used to simulate mixtures of tumor-derived and hematopoietic background DNA²⁰,²¹. Simulations incorporated variation in DNA recovery and assay performance and were implemented using Repast and COPASI⁴³,⁴⁴. Independent simulation runs assessed robustness and supported exploration of multilocus configurations alongside experimental evaluations.

### Statistical analysis

Statistical analyses were performed using GraphPad Prism 11 and R. Data are presented as mean ± s.e.m. unless otherwise indicated. Normality was assessed using the Shapiro–Wilk test, and comparisons used parametric or non-parametric tests appropriate to the distribution and paired or unpaired design. All tests were two-sided. ROC analyses were performed as indicated, with AUCs reported alongside 95% confidence intervals.

Specific tests, sample sizes, replicate handling, exclusion criteria and adjustments for multiple comparisons are described in the corresponding Results and figure legends. Experiments comprising only two independent runs were summarized descriptively without formal statistical testing.

## Data Availability

Processed data supporting the findings of this study are included in the manuscript and its supplementary information. Additional de-identified data generated during the study are available from the corresponding authors upon reasonable request and subject to applicable ethical, privacy and data-use restrictions. Public methylation data used for in silico analyses were obtained from TCGA through the NCI Genomic Data Commons Data Portal. No new sequencing datasets were generated in this study.

https://portal.gdc.cancer.gov/

## Acknowledgements

We thank the participants in the CASCADE and HIDE studies and the clinical, surgical, pathology and laboratory teams involved in participant recruitment, specimen collection and processing, and clinical and histopathological characterization. We acknowledge the investigators and participants who generated and shared the TCGA and Gene Expression Omnibus datasets analyzed in this study. We also thank Fundación Escuela de Medicina Nuclear (FUESMEN) for providing access to laboratory facilities used during the experimental evaluation of Delta-HLD.

## Author contributions

V.B. and E.M.C. conceived and supervised the study. C.C.V., P.A.W. and C.G.S. developed and optimized the assay, performed experiments and analyzed the data. J.S. and Y.A. performed ddPCR experiments and contributed to experimental data generation. L.M. developed and implemented the computational models and simulations. A.R.A. and R.D.M. contributed to experimental work and data analysis. A.L. contributed to project coordination and resources. J.D.R., R.N.M., R.P.R., E.G., C.E.F.D., R.O., G.S., A.C., M.C., W.R.M., G.A.V., P.V. and L.S. contributed to participant recruitment, specimen collection and clinical or histopathological characterization. C.C.V., P.A.W., V.B. and E.M.C. drafted the manuscript. All authors contributed to manuscript review and approved the final version.

## Figure Legends

**Extended Data Figure 1. Development and characterization of the Delta-HLD assay for DNA methylation detection. a,** Schematic showing the matched no-enzyme (NE) and enzyme-treated (E) assay logic. Native DNA undergoes hybridization with adjacent hemi-probes (HP1 and HP2), followed by ligation. In the enzyme-treated (E) reaction, unmethylated targets are cleaved by HhaI and are therefore not amplified, whereas methylated targets are protected from digestion and remain amplifiable. **b,** Schematic of the primer/probe architecture used for Delta-HLD co-reporting, illustrating how multiple target-derived ligation products can share a common amplification and detection system within the same fluorescence channel. **c,** Schematic localization of the SEPTIN9 Delta-HLD target sites relative to the candidate methylation region prioritized in the accompanying discovery study. **d,** Bar plot showing qPCR-based Delta-HLD quantification of SEPTIN9_3520 methylation across six cancer cell lines. Methylation percentages were calculated from matched NE/E reactions corresponding to the same target assessed qualitatively by agarose gel electrophoresis in Fig. 1b. **e,** Representative qPCR amplification profiles for SEPTIN9-based Delta-HLD detection in HT-29 and buffy coat derived DNA. HT-29 DNA is shown in blue and buffy coat DNA in gray; traces with circular markers correspond to no-enzyme (NE) reactions and traces without circular markers correspond to enzyme-treated (E) reactions. **f,** Representative ddPCR analysis of SEPTIN9_3520 Delta-HLD in HT-29 and buffy coat DNA. Left, one-dimensional droplet amplitude plots showing digestion-dependent differences between matched NE and E reactions. Right, corresponding ddPCR concentration estimates for the same conditions. **g,** Schematic of the preliminary plasma cfDNA workflow used for the MIX3 Delta-HLD proof-of-concept, including EDTA blood collection, cfDNA extraction from 1 ml plasma, Delta-HLD processing with the three-locus MIX3 configuration, matched NE/E ddPCR analysis in the FAM channel and methylation readout.

**Extended Data Figure 2. Candidate-panel modeling and artificial liquid-biopsy screening. a,** Box plots showing representative simulated tumor-profile distributions generated for CpG sites 1 and 10 across seven independently sampled tumor-profile sets. The modeled methylation distributions used as input for the COPASI-based panel-size simulations. **b,** Individual simulation curves underlying panel-size optimization showing AUC as a function of the number of co-reported loci across seven independent simulations. **c,** Bar plot showing no-template ddPCR screening of ten candidate 8-locus co-reported panels (8A-8J) used to estimate HP-system background before testing in DNA-containing reactions. Values are shown as copies per μl; bars show mean ± s.e.m. from technical replicates. **d,** Bar plots showing the preliminary qPCR-based Delta-HLD evaluation of the two lowest-background eight-locus configurations, 8A and 8E, in HT-29 DNA versus buffy coat DNA. Bars show methylation levels calculated from matched NE/E reactions, shown as mean ± s.e.m. from three technical replicates. **e,** Artificial liquid-biopsy evaluation of the selected eight-locus configurations. Left and middle box plots showing methylation levels for 8A and 8E in matched control-spiked and tumor-spiked artificial liquid biopsies generated by spiking matched non-tumor or tumor tissue DNA into a constant buffy coat DNA background. Groups were compared using two-tailed exact Wilcoxon matched-pairs signed-rank tests (8A: P = 0.0020; 8E: P = 0.0039; n = 10 pairs per configuration). Right, ROC curve comparing tumor-spiked versus control-spiked artificial liquid biopsies for 8A and 8E (8A: AUC = 0.9100, 95% CI 0.7673 - 1.000, P = 0.0019; 8E: AUC = 0.9300, 95% CI 0.8225 -1.000, P = 0.0012; n = 10 control-spiked and n = 10 tumor-spiked artificial liquid biopsies per configuration). **f,** Evaluation of 5-locus derivatives from the 8A and 8E configurations. Left, box plots showing methylation levels in matched control-spiked and tumor-spiked artificial liquid biopsies for 8A-5C, 8A-5E, 8E-5A and 8E-5J. Groups were compared using two-tailed exact Wilcoxon matched-pairs signed-rank tests (8A-5C: P = 0.0039; 8A-5E: P = 0.0195; 8E-5A: P = 0.059; 8E-5J: P = 0.0020; n = 10 pairs per subset). Right, ROC curve comparing tumor-spiked versus control-spiked artificial liquid biopsies for each five-locus subset (8A-5C: AUC = 0.9700, 95% CI 0.9019 - 1.000, P = 0.0004; 8A-5E: AUC = 0.8700, 95% CI 0.6909 - 1.000, P = 0.0052; 8E-5A: AUC = 0.7400, 95% CI 0.5199 - 0.9601, P = 0.0696; 8E-5J: AUC = 0.9600, 95% CI 0.8747 - 1.000, P = 0.0005; n = 10 control-spiked and n = 10 tumor-spiked artificial liquid biopsies per subset). **g,** Performance of the original three-locus MIX3 configuration under the same qPCR-based artificial liquid-biopsy conditions used for model-guided panel screening. Left, box plot showing methylation levels in matched control-spiked and tumor-spiked artificial liquid biopsies; groups were compared using a two-tailed exact Wilcoxon matched-pairs signed-rank test (P = 0.0020; n = 10 pairs). Right, ROC analysis for tumor-spiked sample discrimination (AUC = 0.9000, 95% CI 0.7560 - 1.000, P = 0.0025; n = 10 control-spiked and n = 10 tumor-spiked artificial liquid biopsies). **h,i,** Direct paired comparison of MIX3 and the optimized 8E-5J configuration in eight independent tumor-spiked artificial liquid-biopsy samples. **h,** Box plot showing the summary comparison; groups were compared using a two-tailed exact Wilcoxon matched-pairs signed-rank test (P = 0.0044; n = 8 paired samples). **i,** Paired plot showing the same samples, with lines connecting matched MIX3 and 8E-5J measurements, showing consistently higher methylation values with 8E-5J.

**Extended Data Figure 3. Optimization of digestion-control concentration and representative triplex detection profiles. a,** Dot plots showing qPCR-based evaluation of the digestion-control (DC) titration. Left, enzyme-treated/no-enzyme (E/NE) signal ratios across DC concentrations (0.015, 0.0015 and 0.0003 pg/µl). Right, corresponding Cq values for matched NE and E reactions, showing the expected digestion-dependent Cq shift after enzyme treatment. Individual points represent technical replicates (n = 3 per condition). **b,** Dot plot showing ddPCR-based evaluation of the synthetic DC template across the same three concentrations. E/NE signal ratios are shown across DC concentrations to identify a low DC input compatible with reliable detection and efficient digestion monitoring. Individual points represent technical replicates. **c,** Representative ddPCR one-dimensional amplitude plots for FAM and HEX channels in healthy-donor buffy coat-derived genomic DNA and HT-29 DNA under matched NE and E conditions. FAM reports the 8E-5J target panel, whereas HEX reports the synthetic DC. **d,** Plots showing corresponding ddPCR concentration estimates from the conditions shown in c, illustrating FAM and HEX detection across buffy coat DNA and HT-29 DNA in matched NE/E reactions. **e,** Representative qPCR amplification profiles for the final triplex configuration. Across all channels, solid traces correspond to HT-29 DNA and traces with circular markers correspond to buffy coat DNA. In the FAM channel, the 8E-5J target panel is shown for matched NE and E reactions, with NE and E traces displayed in dark blue and cyan, respectively. In the HEX channel, DC is shown in matched NE and E reactions, with the undigested NE fraction in orange and the enzyme-treated E fraction in yellow, illustrating the expected digestion-dependent delay. In the Cy5 channel, the amplification control (AC) is shown in matched NE and E reactions, with NE in violet and E in pink, illustrating comparable amplification across the matched pair. Horizontal lines indicate the fluorescence threshold used for Cq determination.

**Extended Data Figure 4. Preanalytical optimization and analytical operating range for the final qPCR Delta-HLD workflow. a,** Bar plot showing the effect of plasma input volume on cfDNA recovery. cfDNA concentration is shown for extractions performed from 1.5 ml and 3 ml plasma, supporting the use of increased plasma input in the optimized workflow. **b,** Bar plot showing the effect of blood-collection tube type on cfDNA recovery. cfDNA concentration is shown for plasma collected in EDTA and Streck tubes under matched processing conditions. **c,** Dot plot comparing cfDNA extraction workflows. Cq values for the no-enzyme (NE) FAM reaction are shown after vacuum-based and magnetic bead-based extraction, supporting transition to the bead-based workflow used for downstream plasma evaluation. **d,** Input-range plot showing analytical evaluation for the final triplex qPCR workflow across serial unmethylated DNA inputs. 8E-5J co-reported methylation signal, FAM NE Cq values, HEX digestion-control behavior and Cy5 amplification-control Cq values are shown across the dilution series. FAM NE Cq values increased approximately linearly with decreasing DNA input up to Cq 35, which was defined as the upper input-acceptance limit for downstream plasma analysis. Control-channel behavior remained stable across the tested range. The arrow indicates 1.56 ng, the conservative unmethylated DNA input selected as the fixed background for subsequent methylated-DNA spike-in experiments. **e,** Bar plot showing spike-in titration of methylated control DNA into a fixed 1.56 ng unmethylated background. 8E-5J co-reported methylation signal decreased with decreasing methylated input; the arrow indicates 0.18 ng, the lowest methylated input yielding a reproducibly detectable signal under these conditions. **f,** Bar plot showing independent dilution of HT-29 DNA into buffy coat DNA, showing concordant reduction in 8E-5J Delta-HLD methylation signal across the low-input range.

**Extended Data Figure 5. Target-level evaluation of HCC-associated methylation targets.** Heat map showing qPCR-based Delta-HLD methylation levels for candidate HCC targets in cirrhotic liver tissue specimens (n = 12) and HCC tumor specimens (n = 11). Rows correspond to individual methylation targets and columns to individual tissue specimens; color intensity indicates methylation level. The same tissue cohort was used for locus-specific experimental evaluation of six targets in the accompanying discovery study.

**Extended Data Figure 6. Screening of Delta-HLD reaction schemes for reduced genomic DNA recognition and preservation of short control-template detection. a,** Schematic showing the three Delta-HLD reaction schemes tested. P1 corresponds to the original Delta-HLD protocol. P2 eliminates the initial 5-min high-temperature DNA denaturation and the separate 25 °C incubation after probe addition, whereas P3 retains the P1 sequence but reduces both high-temperature steps to 80 °C. P2 was selected for subsequent analyses based on reduced genomic DNA recognition while preserving detection of short control templates. **b,** Dot plots showing Cq comparative evaluation of P1-P3 across FAM, HEX and Cy5 channels. Genomic HT-29 DNA was used to evaluate FAM panel-associated genomic DNA detection, whereas short synthetic digestion-control (DC) and amplification-control (AC) templates were used to monitor preservation of short control-template detection in HEX and Cy5. Cq values are shown for matched no-enzyme (NE) and enzyme-treated (E) reactions. Compared with the original P1 condition, P2 reduced genomic HT-29 DNA detection in FAM while preserving short-template detection in HEX and Cy5, supporting its selection for subsequent platelet-retaining protocol analyses.

