## Supplementary figures and images for "Panel-level multilocus methylation quantification in native cell-free DNA by PCR-compatible sequential enzymatic processing"

### Extended Data Fig 1

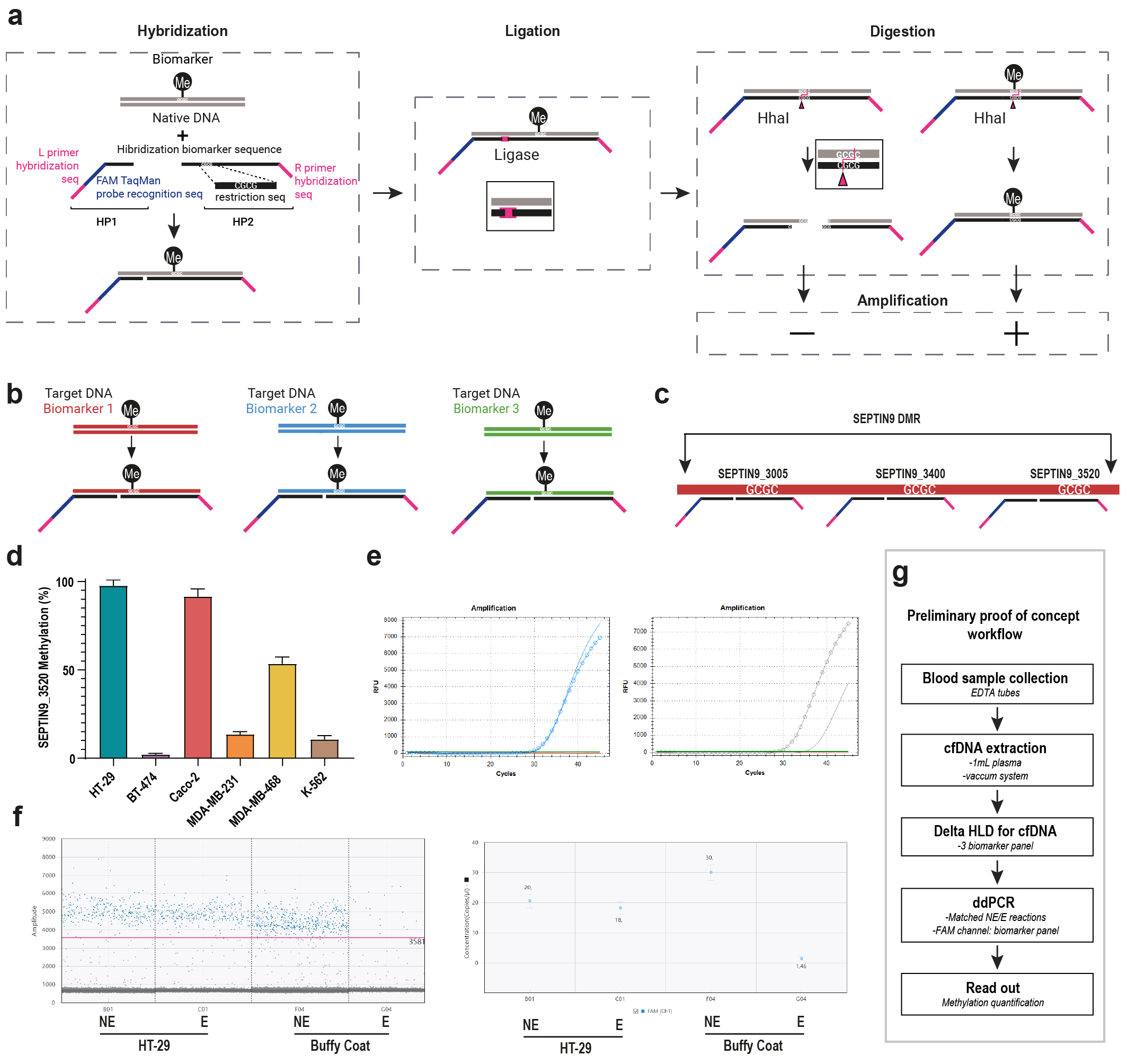

### Extended Data Fig 2

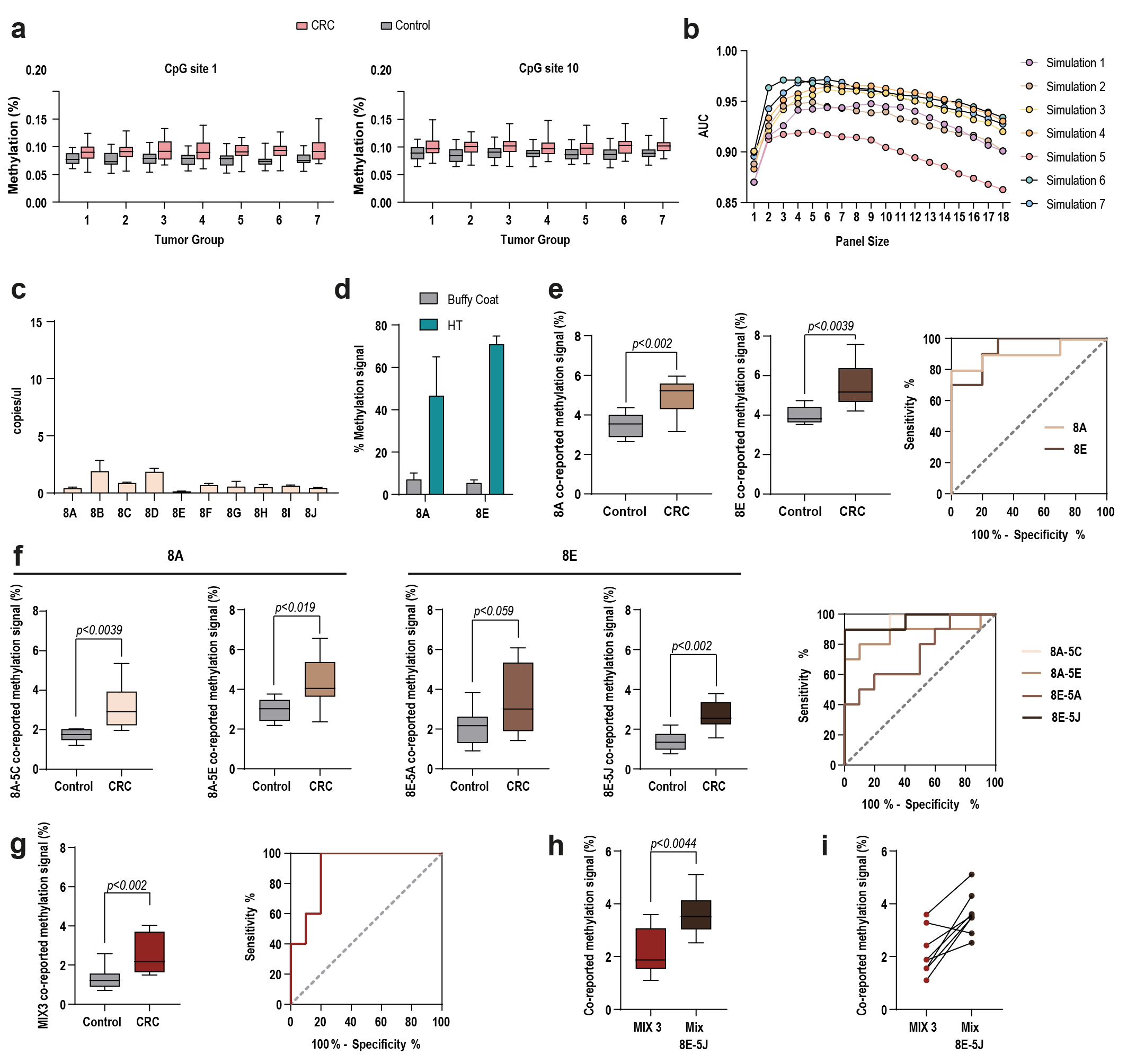

### Extended Data Fig 3

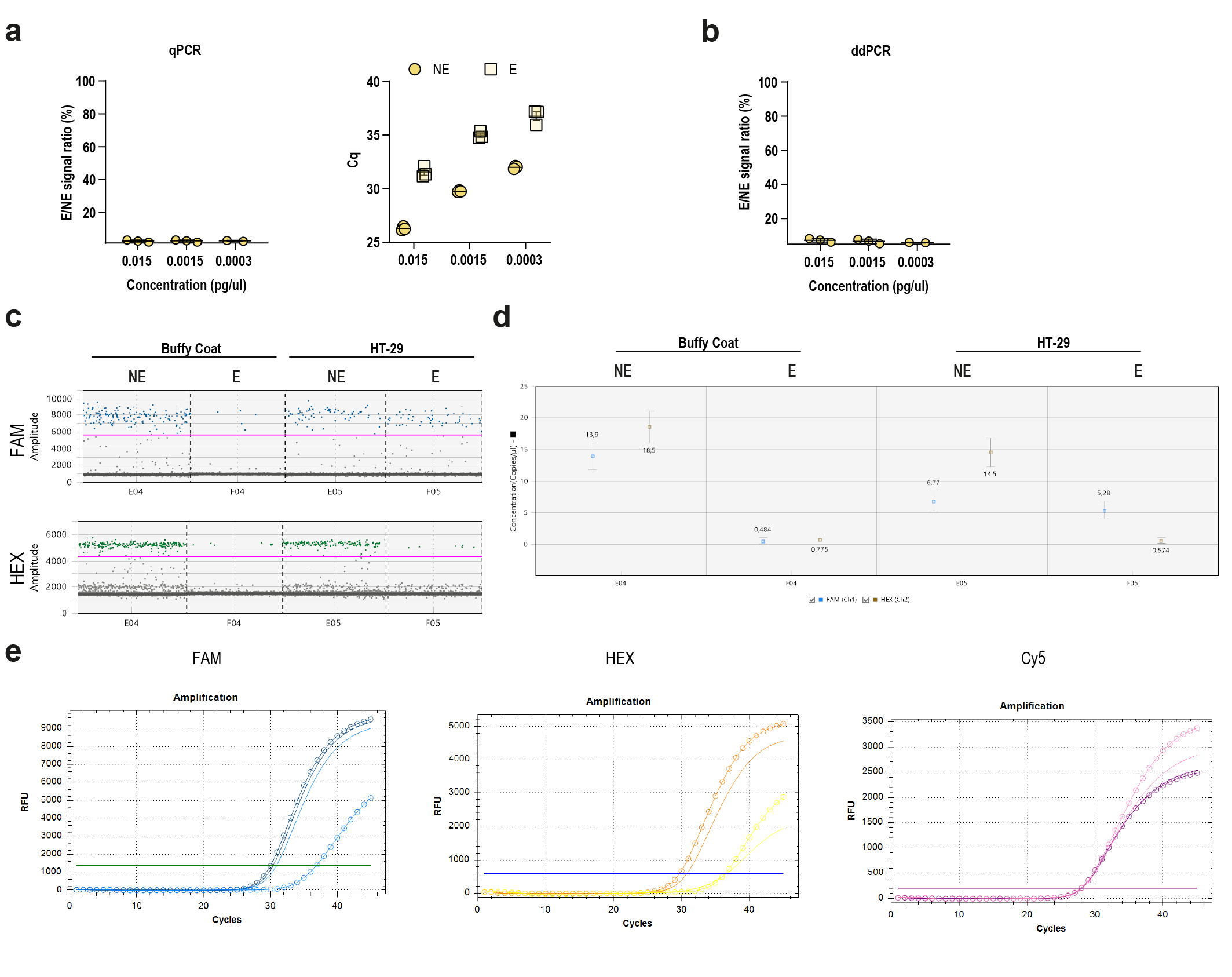

### Extended Data Fig 4

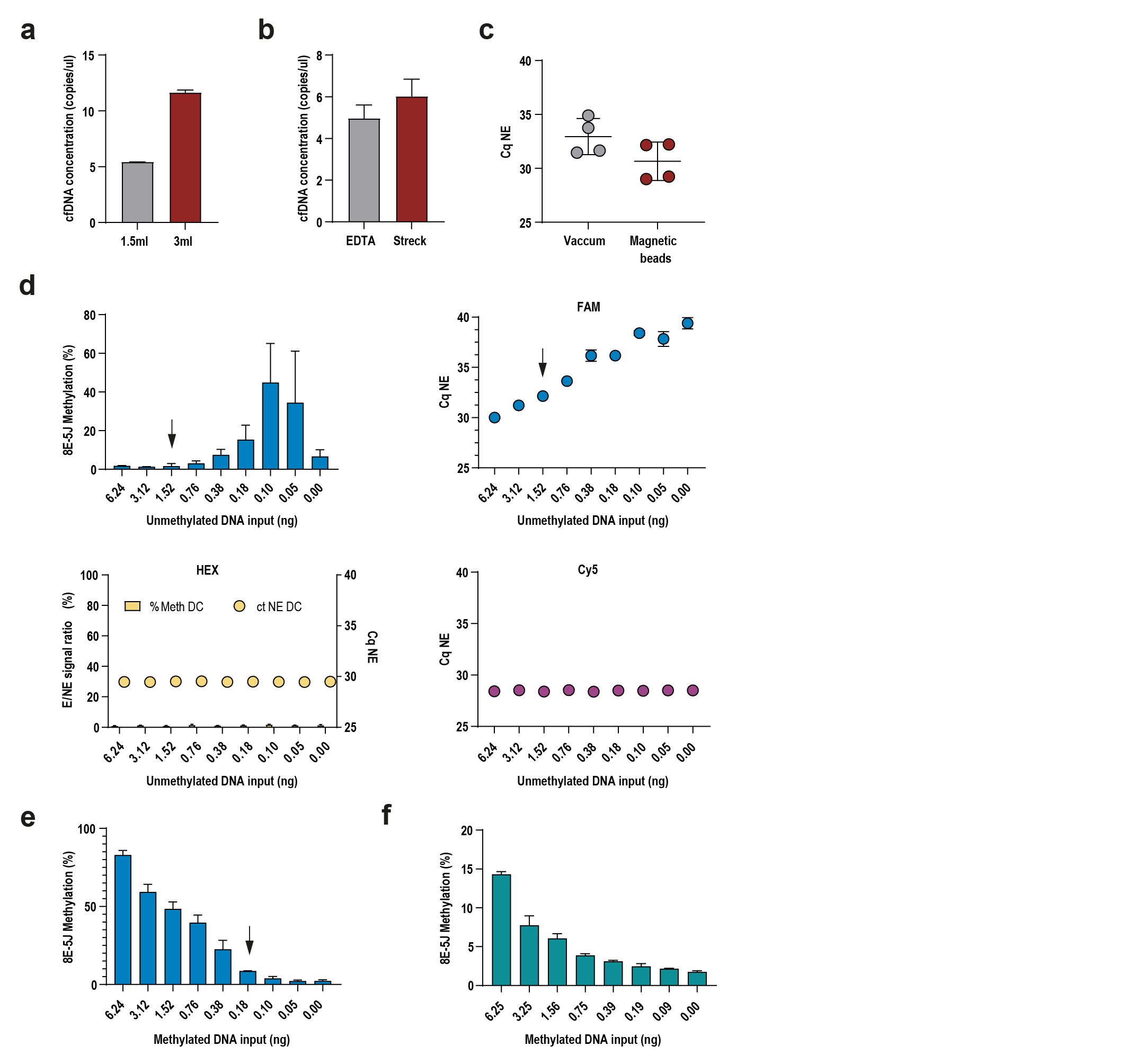

### Extended Data Fig 5

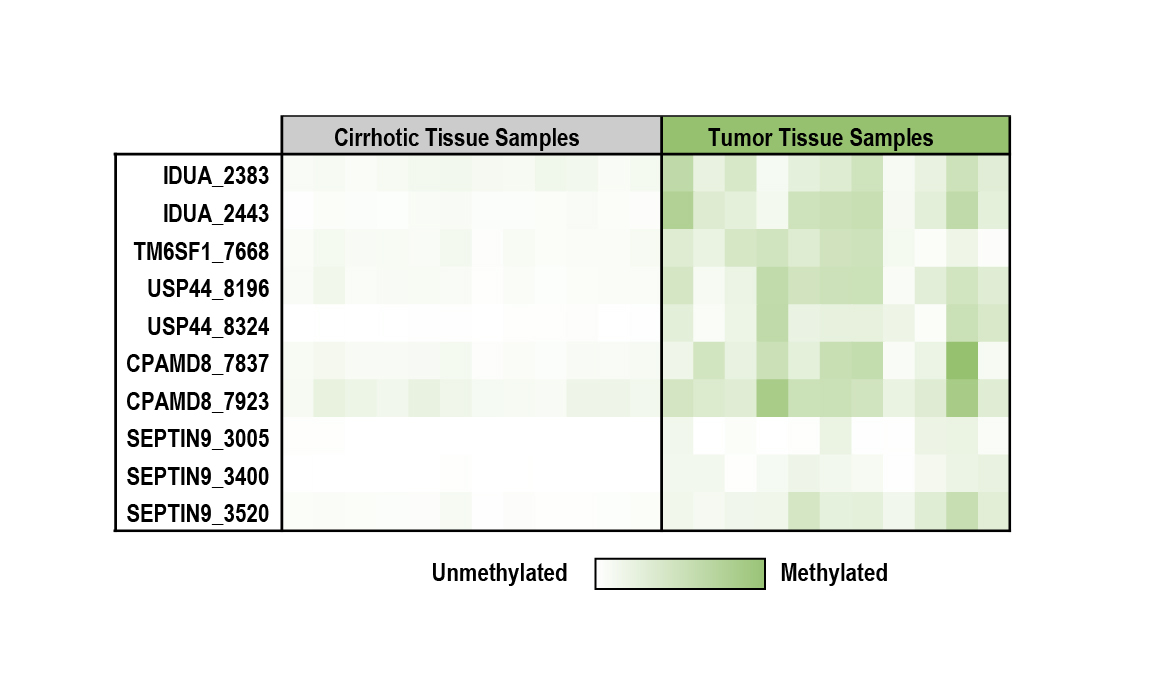

### Extended Data Fig 6

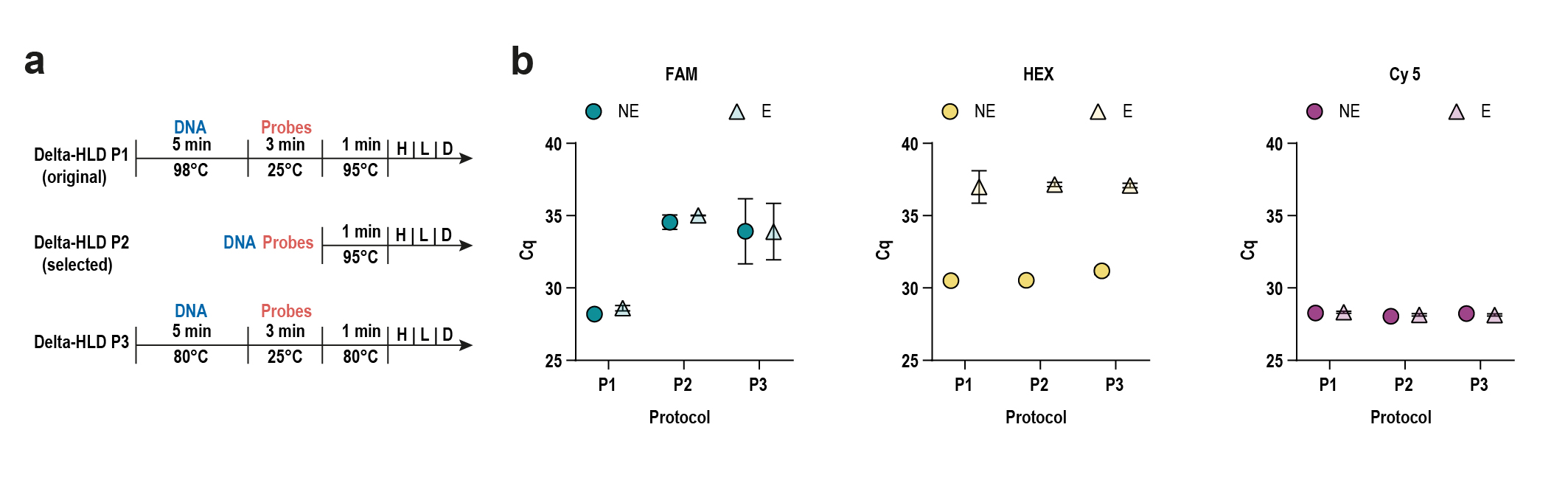
